# *Celeste*: A cloud-based genomics infrastructure with variant-calling pipeline suited for population-scale sequencing projects

**DOI:** 10.1101/2025.04.29.25326690

**Authors:** Noora Siddiqui, Breanna Lee, Victoria Yi, Jesse Farek, Ziad Khan, Sara E Kalla, Qiaoyan Wang, Kimberly Walker, James Meldrim, Christopher Kachulis, Michael Gatzen, Niall J. Lennon, Shyamal Mehtalia, Severine Catreux, Rami Mehio, Richard A Gibbs, Eric Venner

**Author notes:** Corresponding author: Eric Venner.

## Abstract

**Background:** The *All of Us* Research Program (*All of Us*) is one of the world’s largest sequencing efforts that will generate genetic data for over one million individuals from diverse backgrounds. This historic megaproject will create novel research platforms that integrate an unprecedented amount of genetic data with longitudinal health information. Here, we describe the design of *Celeste*, a resilient, open-source cloud architecture for implementing genomics workflows that has successfully analyzed petabytes of participant genomic information for *All of Us* – thereby enabling other large-scale sequencing efforts with a comprehensive set of tools to power analysis. The *Celeste* infrastructure is tremendously scalable and has routinely processed fluctuating workloads of up to 9,000 whole-genome sequencing (WGS) samples for *All of Us*, monthly. It also lends itself to multiple projects. Serverless technology and container orchestration form the basis of *Celeste*’s system for managing this volume of data.

**Results:** In 12 months of production (within a single Amazon Web Services (AWS) Region), around 200 million serverless functions and over 20 million messages coordinated the analysis of 1.8 million bioinformatics, quality control, and clinical reporting jobs. Adapting WGS analysis to clinical projects requires adaptation of variant-calling methods to enrich the reliable detection of variants with known clinical importance. Thus, we also share the process by which we tuned the variant-calling pipeline in use by the multiple genome centers supporting *All of Us* to maximize precision and accuracy for low fraction variant calls with clinical significance.

**Conclusions:** When combined with hardware-accelerated implementations for genomic analysis, Celeste had far-reaching, positive implications for turn-around time, dynamic scalability, security, and storage of analysis for one hundred-thousand whole-genome samples and counting. Other groups may align their sequencing workflows to this harmonized pipeline standard, included within the *Celeste* framework, to meet clinical requisites for population-scale sequencing efforts. *Celeste* is available as an Amazon Web Services (AWS) deployment in GitHub, and includes command-line parameters and software containers.

## Background

As the field of genomic medicine continues to grow, diversity and inclusion within large scale population projects are imperative to ensure adequate scientific progress in addressing health disparities ^1,2^. Population-scale studies that span multiple ancestries can improve the power of genome wide association studies (GWAS), pharmacogenomics, and other genomic analyses to diagnose, monitor, and treat disease ^3–5^.

However, population-scale projects are computationally intensive and require secure, bulk storage of data along with scalability of infrastructure. *All of Us* poses two additional challenges: the harmonization of workflows across several sequencing centers, and the requirement for analysis of data in a clinical environment that meets the standards set by the US Food and Drug Administration (FDA) ^6^.

### All of Us Research Program

*All of Us* is designed to blend genetic data with health information for a diverse cohort of one million participants, thereby creating a research resource with data centered around individuals who have not been traditionally included in biomedical research ^7,8^. This program, powered by a longitudinal group of geographically, demographically, and medically diverse individuals, is intended to enable a wide variety of ongoing scientific innovation at the forefront of precision medicine ^8^. Select genomics data, notably secondary findings in American College of Medical Genetics and Genomics (ACMG) genes and pharmacogenomics information, are returned directly to participants who elect to receive this information ^9^. The data generation and results workflow for *All of Us* are conducted under an investigational device exemption authorized by the U.S. Food and Drug Administration (FDA) ^6^.

Three centers support the genome sequencing component for this project; these are the Baylor-Hopkins Clinical Genomics Center, the Broad Institute of MIT and Harvard, and the Northwest Genomics Center at the University of Washington. Once data is sequenced and analyzed at one of these three sequencing centers, it is sent to the *All of Us* Data and Research Center (DRC) at Vanderbilt University, where it is compiled into the Researcher Workbench^10^–a cloud-based research platform available for registered users.

### Challenges of Scale and Cloud Migration

The scale of *All of Us* is momentous. While other projects have sequenced sizable cohorts, returned genomic findings to participants, or created rich data resources ^11–14^, *All of Us* has positioned itself uniquely in terms of all three of those above aims with the help of 1 million ancestrally and medically diverse participants.^7^

The standard WGS pipeline entails 30 different quality control, bioinformatic analysis, and clinical reporting jobs, producing over 300 files and over 150Gb of transformed data per sample. Processing this amount of clinical data requires a considerable investment in acquiring and maintaining erratically used computational resources. As genomics analysis research and clinical analysis requirements persistently evolve, the burden of accurately predicting the number of compute and storage servers needed for a particular project is immense. Further, these genomics analysis workloads are fundamentally bursty, a use-case that lends itself to a cloud environment.

There have been many approaches to scalable computing for genomic projects ^15–17^, but our infrastructure as code deployment, packaged with containerized software components and specific command line parameters can quickly be modified and utilized to take advantage of either the workflow orchestration component, or the entire sample processing pipeline. To account for the All of Us WGS analysis workload, we focused on decreasing sample processing time and improving storage modernization, scalability, security, and cost efficiency, via a dynamic Infrastructure as a Solution (IaaS) cloud solution.

Our team migrated to a hybrid-cloud architecture and engineered *Celeste* – a highly available, automated, and scalable clinical genomics analysis deployment with the flexibility to handle bursty workloads in a cost-effective and secure manner ^18^. *Celeste* was deployed at-scale within 3 months and now processes high-throughput human whole genome workloads for *All of Us* ^19^.

|  |  |
| --- | --- |
| **Problem:** | The absence of a scalable, secure, open-source cloud-based infrastructure to handle the massive amount of genomic data generated by large-scale sequencing efforts. |
| **What is Already Known:** | Reliable detection of clinically significant variants at population scale requires an adjusted approach from typical workflows. Potential mosaic variants are of research interest and should not be overlooked in population datasets. Lastly, bioinformatics pipelines lack standardization and rely on functional equivalence. |
| **What This Paper Adds** | Here, we define and open-source an extensible infrastructure suitable for population scale sequencing programs and document a harmonized pipeline tailored for that purpose. |

## Implementation

### Cloud-based Architecture

*Celeste* is a robust infrastructure deployment that provisions an Amazon Virtual Private Cloud (VPC) with the needed networking components to process clinical samples securely, along with an AWS Batch job management system, a workflow orchestration system that comprises of serverless AWS Lambda functions and Amazon Simple Notification Service (SNS) topics that respond to the appearance of specific data files, multiple software containers for jobs that carry out standardized bioinformatics pipeline steps, a CloudWatch EventBridge and serverless system that automatically handles job failures and resubmissions due to Spot Instance interruption, and S3 lifecycle rules that automatically handle archival and cleanup for pipeline outputs. This infrastructure is captured in CloudFormation templates, complete with Lambda functions containing specific bioinformatic software command-line parameters and a data- tagging system for use in lifecycle management. The bioinformatics pipeline packaged within *Celeste* is suitable for clinical and population-scale sequencing projects. The reusability of pipeline components allows this. The CloudFormation management and GitHub repository for *Celeste* enables re-deployment and facilitates reuse of the infrastructure across multiple clinical sequencing projects in addition to *All of Us*.

A maximum of 400,000 jobs flow through Celeste monthly. To handle the burst workloads typical of next-generation sequencing workflows, we developed an AWS Batch job management system that allows us to provision thousands of instances, scale servers dynamically depending on demand, and utilize Spot instances when possible, for cost savings. Workflow orchestration within the cloud infrastructure is decoupled and event driven. New sample data in Amazon S3 triggers an SNS message, which catalyzes a cascade of serverless AWS Lambda functions. These Lambdas identify the file type and submit relevant AWS Batch jobs depending on both S3 prefix and file extension. Additional Lambdas work in concert with S3 lifecycle management rules to automate data archival in accordance with compliance protocols. (See Methods).

**Figure 1.**
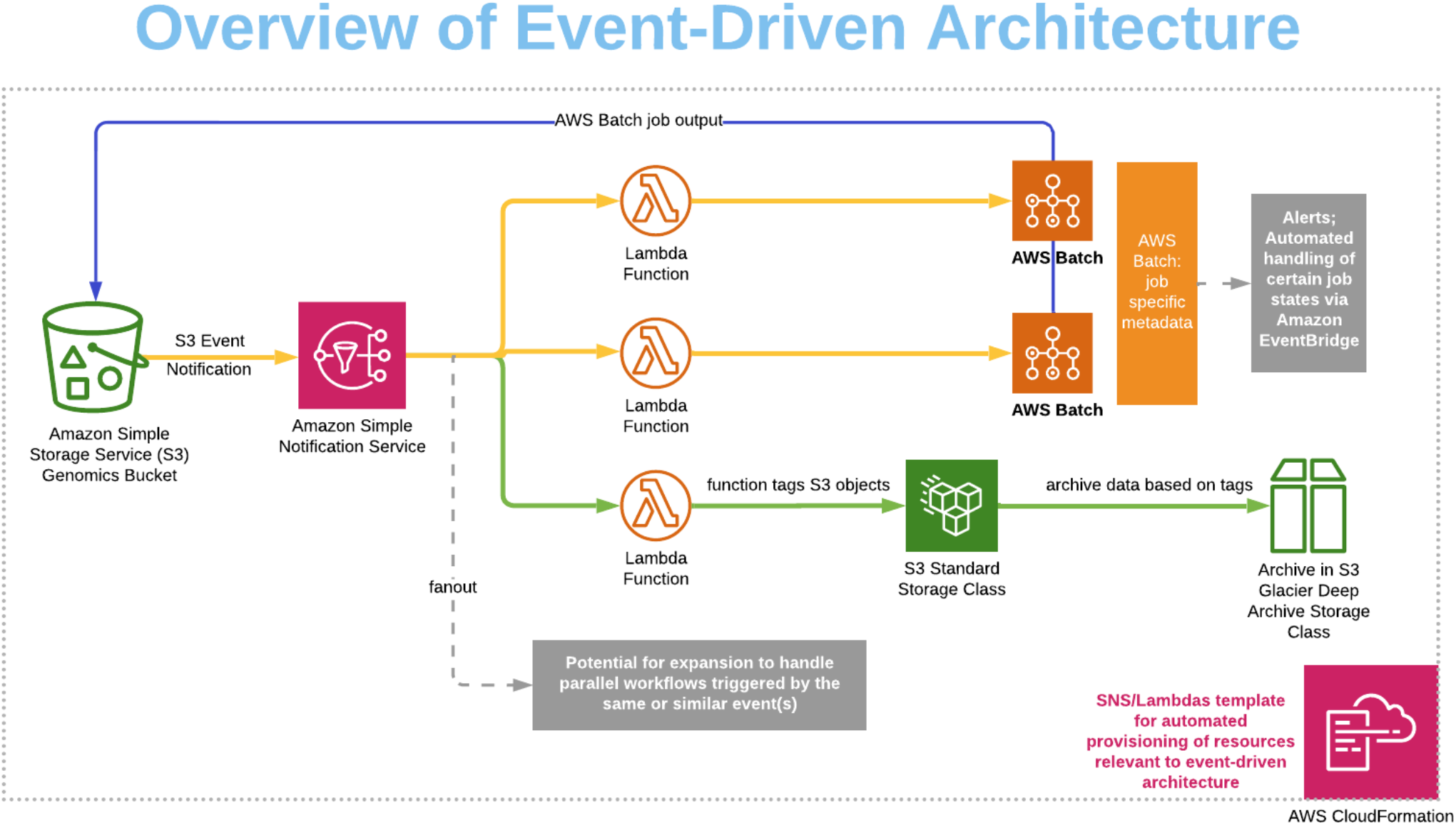
Overview of architecture. Celeste is a flexible, event-driven architecture that allows for the implementation of multiple clinical genomics workflows. After an upload from local storage, An S3 Event Notification starts the workflow. The Amazon Simple Notification Service (SNS) acts as a pub/sub service to decouple and scale microservices, distributed systems, and serverless applications. Here it is used to trigger one or more Lambda functions, fanning-out jobs for parallel processing. Bioinformatics pipelines are implemented on AWS Batch, using job-specific metadata. EventBridge alerts aid in monitoring pipeline operations. The data lifecycle is automated, with large files aging out into S3 Glacier Deep Archive automatically. All elements of the system are captured in CloudFormation templates.

We modularized our architecture by containerizing applications using Docker. A wrapper script handles conditional input and output to and from S3 when provided with S3URI command-line parameters, allowing jobs to be utilized identically on-premise and within AWS Batch.

We utilize tools within AWS CloudWatch to track logs and metrics from the deployed infrastructure solution. By combining log insights with comprehensive dashboards and alarms, we effortlessly investigate and troubleshoot system disruptions across services. Amazon EventBridge rules, in conjunction with Lambda, allow us to automatically handle job failures and resubmissions. Sample tracking occurs outside of the *Celeste* deployment via AWS Athena and Amazon QuickSight.

Architecture deployments can span from 1-4 Availability Zones (AZs) with 2 recommended as a minimum for high availability. Outbound internet access for private AWS Batch instances is enabled via Network Address Translation (NAT) Gateways in public subnets. This system negates any public inbound internet connection to analysis jobs, while allowing certain software to send usage information to licensing servers. For the 4 AZ deployment, 2 NAT gateways form a sufficient, highly available system to manage the negligible amount of outbound internet access required by our sequencing analysis workflow. Multiple AZs provide the foundation for horizontal scalability, enable robust disaster recovery strategies, and can provide cost optimizations.

**Figure 2.**
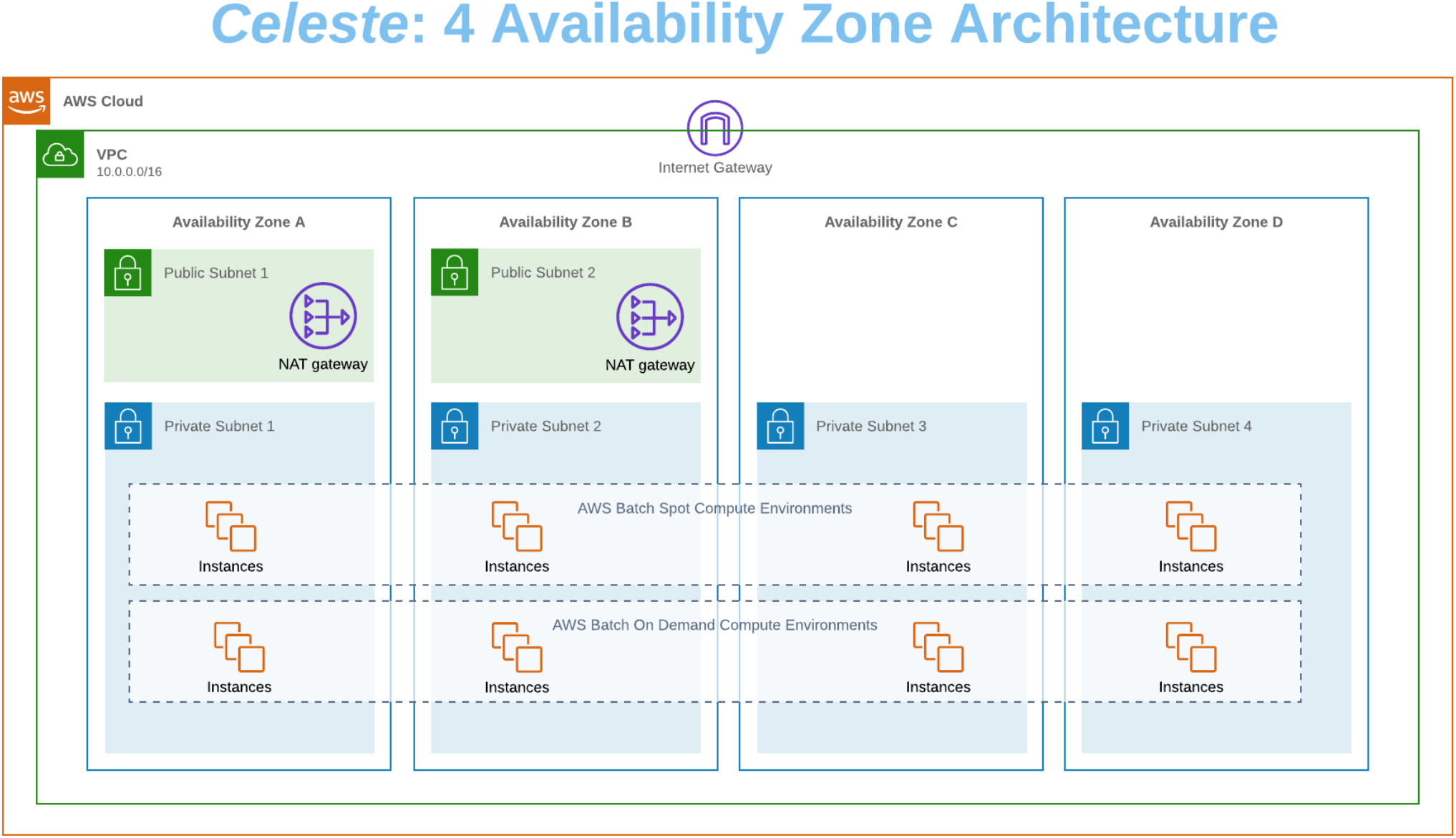
Use of multiple availability zones increases the resilience of the Celeste infrastructure. Building infrastructure across multiple availability zones (AZs) offers increased availability, improved fault tolerance, can promote scalability, and allow for simpler disaster recovery. It ensures that our pipeline remains available even if one AZ experiences an outage or becomes inaccessible.

**Figure 3.**
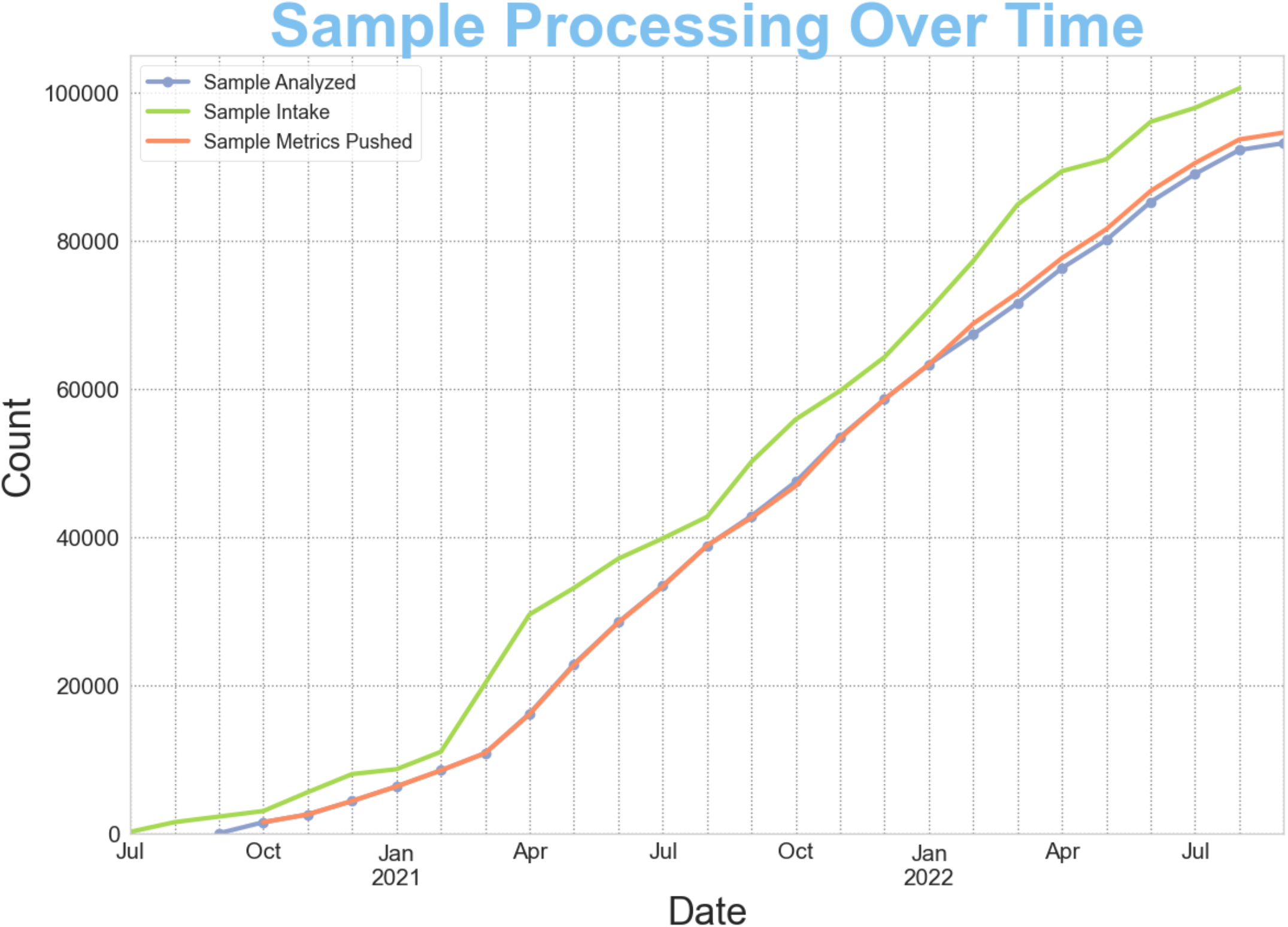
Cumulative samples processed through Celeste. The system has been used to analyze over 360,000 WGS reprocessing samples and 115,000 production WGS samples. Its scalability allowed for a ramp-up in April of 2021 and has kept pace with sample submission and data production.

### Speed of Sample Processing

Using the agility of Celeste, we undertook a major reprocessing effort in which we analyzed over 32 thousand whole genome samples in under 20 days (Table 1). We rapidly provisioned reprocessing resources, which mirrored deployments of Celeste across two AWS Regions. Each deployment takes less than 45 minutes. We then unarchived the needed internal samples that had entered the S3 Glacier Deep Archive and downloaded CRAM files via the DRC’s

Google Cloud Platform bucket into S3. The resulting 32,419 WGS samples ran through our cloud-based infrastructure for bioinformatic analysis in 17 days. The rate limiting step of this project was the availability of field-programmable gate array (FPGA) instances due to increased demand for those resources among AWS customers. Our multi-regional approach addressed this challenge and allowed us to reprocess samples at a steady rate without the need to reserve instance capacity. Subsequently, we have taken on a further reprocessing effort to unify the *All of Us* research program’s data onto the DRAGEN 3.7.8 version, completing over 330,000 whole genome analyses in less than 6 months (Figure 4b).

**Figure 4.**
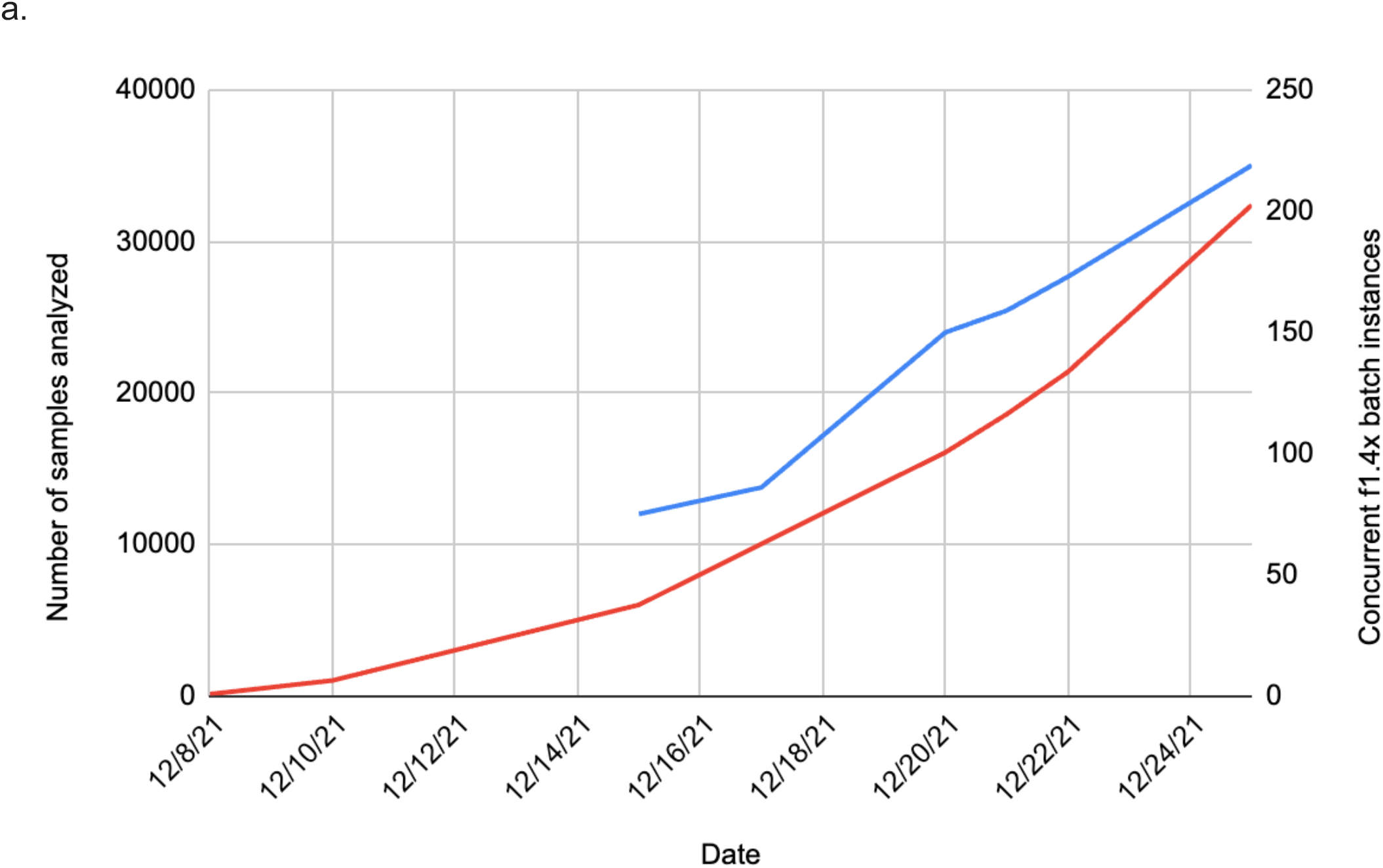
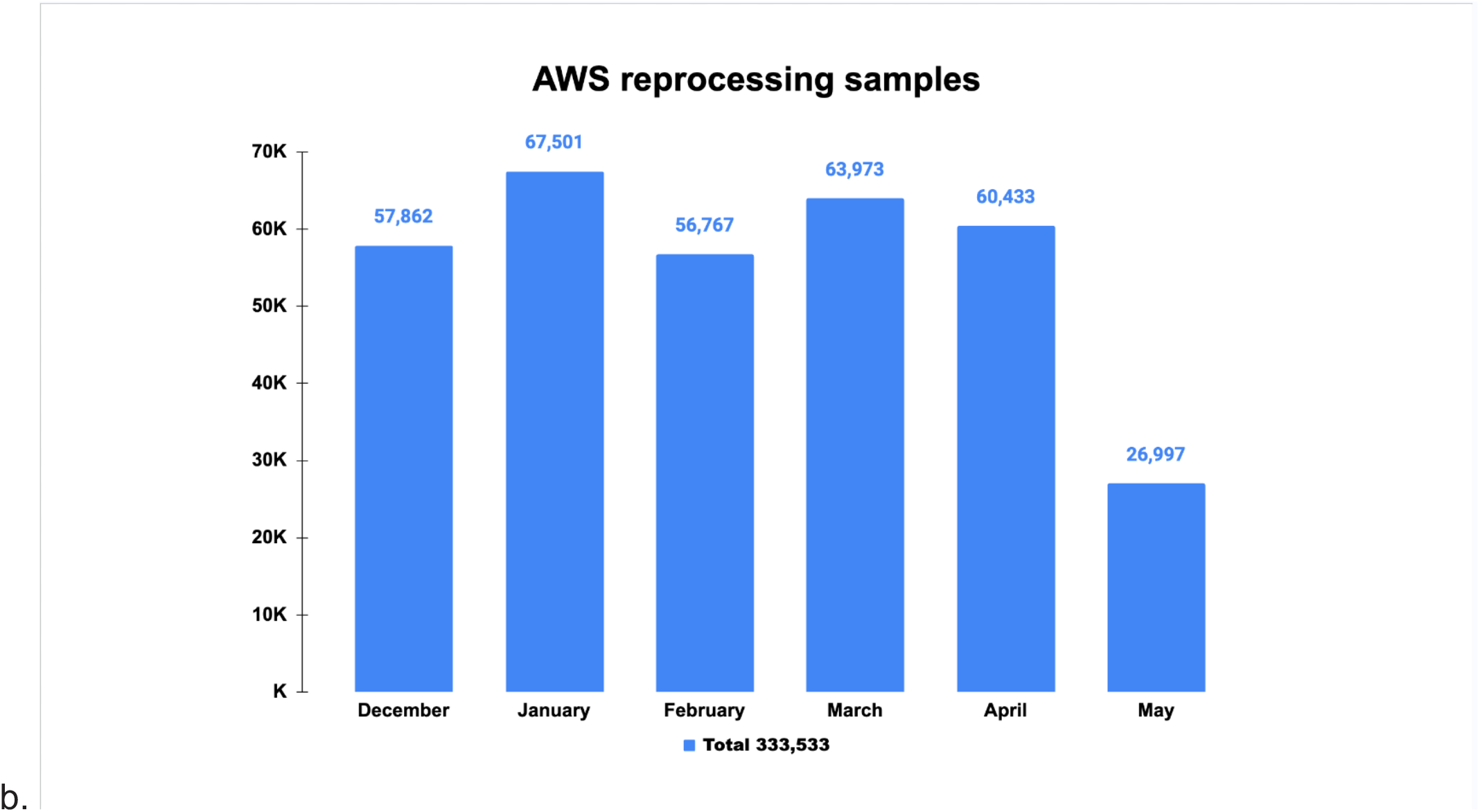
All of Us Reprocessing efforts. 4a. First, this infrastructure was used to carry out a large-scale reprocessing workload in which 32,419 whole genome samples were processed in 17 days. Subsequently, after a program-wide move to Dragen 3.7.8, this infrastructure was used to reprocess 333,533 samples over the course of 6 months (4b).

### Storage Modernization

WGS workflows necessitate managing large volumes of data, and the ease of data lifecycle management is a major advantage of the Celeste infrastructure. Along with these features and the use of multipart uploads in our data transfers, we have also leveraged a unique cloud solution, S3fs. This feature behaves as a network attached drive and allows us to treat an S3 bucket (object storage) as a local EC2 file system via a specified mount point. This process results in increased space, memory, and time efficiency when running AWS Batch jobs that require access to large files in S3. These large files can be accessed without having to download them directly onto the running compute node. In our clinical pipeline, we’ve implemented the S3fs solution to run our in-house variant annotation tool (Cassandra) on EC2 instances. Given Annovar output, Cassandra references multiple database files stored in an S3 bucket and accessed via S3fs, thus decreasing runtime and improving cost efficiency.

### Harmonization and Benchmarking the DRAGEN BioIT Platform

The first bioinformatics pipeline implemented on Celeste employed Illumina’s Dynamic Read Analysis for GENomics (DRAGEN) Bio-IT platform ^20^, which provides hardware (FPGA) accelerated workflows for genomics analyses and generates a variety of metrics for assessing quality at several stages of the secondary analysis workflow. Previous benchmarks, including the Precision FDA challenges, illustrated the speed and accuracy of DRAGEN ^21,22^. A full description of the steps included in this pipeline can be found in Methods.

For the FDA’s Investigational Device Exemption process, we examined performance of this pipeline on 1,210 validation samples^6^. Our in-house HgV Mercury ^15^ pipeline with the variant calling software, xAtlas ^23^, took a mean run time of 78.86 wall-clock hours for a 30x human whole genome. DRAGEN took a mean wall-clock time of 1.40 hours, making it over 56 times faster.

As part of our work readying this pipeline, we tested sample preparation, sequencing, and primary and secondary bioinformatics performance on a control sample designed with variants of known low allelic fractions. Low allele fraction variants are an active area of research, due to their potential importance in disease through either chromosomal hematopoiesis of indeterminate potential (CHIP) or mosaicism ^24–28^. However, many variant calling programs are not optimized to detect these variants and they can be systematically removed from large datasets. By default, DRAGEN parameters are optimized towards a maximum f-measure, with sensitivity and precision given equal weight. This default biases against the detection of low allele fraction variants since many of these variant calls have a higher chance of being false positive as they skew away from the expected diploid model that yields a 50% allele fraction for heterozygous variants. The solution to this problem was to tune the DRAGEN parameters to improve the detection of low allele fraction variant calls, while minimizing the degradation in precision and f-measure. The initial search was done using DRAGEN version 3.3.7, and subsequent version changes were evaluated on this control sample to ensure performance is maintained.

As a result of this search, we introduced two non-default parameters to the DRAGEN command line (--vc-min-call-qual=5, --vc-frd-max-effective-depth=40). Since low allele fraction calls tend to have a lower confidence than calls with allele fraction closer to 50%, the setting for the QUAL confidence threshold (--vc-min-call-qual) is lowered from the default value of 10 to 5 to successfully detect these variants (AoU Adopted Parameters in Table 2). In addition, inside the DRAGEN variant caller, the confidence of heterozygous variants gets lowered by a penalty value that is a function of depth and observed allele fraction. The penalty increases as the allele fraction decreases away from the expected diploid (50%) allele fraction, and increases with depth as well. The default parameter value allows for an unconstrained penalty, which we limit by setting --vc-frd-max-effective-depth=40 to help in the detection of skewed allele fraction calls. Although not the version harmonized for cross-program use and not reviewed by the *All of Us* program, DRAGEN v4.3 (releasing in Q2’24) introduces a specific mosaic calling mode.

**Table 2.** Parameter tuning on the Tru-Q 1 (5% Tier) Reference Standard control sample. Initial benchmarking on a specialized control sample raised concerns about sensitivity for variants with skewed allele fractions, which may be of research interest for user’s of the All of Us program’s data. The DRAGEN team worked to tune parameters to improve the detection of this class of variants, while minimizing the degradation of precision and f-measure.

| Chromosome | Genomic Position | Reference | Alternate | Expected Allelic Frequency (%) | DRAGEN 3.3.7 Default Parameters (FRD ON, QUAL=10) | AoU Adopted DRAGEN Parameters |
| --- | --- | --- | --- | --- | --- | --- |
| 3 | 178952085 | A | G | 30 | ✓ | ✓ |
| 12 | 25398281 | C | T | 25 | ✗ | ✓ |
| 7 | 55241707 | G | A | 16.7 | ✗ | ✓ |
| 7 | 140453136 | A | T | 8 | ✓ | ✓ |
| 12 | 25398284 | C | G | 5 | ✗ | ✓ |

All three sequencing centers opted to take advantage of DRAGEN’s FPGA accelerated analysis and utilize the parameter set resulting from our adjustments as the analysis standard across the *All of Us*, presenting the opportunity presented to completely harmonize our distinct bioinformatics pipelines at the command-line level, rather than pursuing functional equivalence^29^. The program harmonized on DRAGEN version 3.7.8. The advantages to this approach were many. Importantly, the genome centers were able to take advantage of validation data generated by other sites, including for clinical samples where the participant data itself could not be shared^6^ . Further, harmonization simplified concordance calculations, decision making, and development. For example, instead of creating unique but compatible thresholds for variant calls, the genome centers were able to generate a single set of thresholds for key metrics. The process by which we tested the same control samples and insisted upon exact equivalence for results across all three centers also has far-reaching, positive implications for data consistency and integrity for the *All of Us*.

## Results and Discussion

Early cloud workloads began with over 1,000 whole genomes per month. As of mid-2022, Celeste has handled a maximum of 9,241 WGS samples in a single month for *All of Us*, automatically scaling from 0 to thousands of concurrently running EC2 instances to handle burst workloads. Samples that have completed bioinformatic analysis keep pace with sample intake for a cumulative sum of close to 100,000 WGS *All of Us* samples and counting.

To date, over 7 petabytes and 40,000,000 files of transformed data were successfully processed through the pipeline, all within a single AWS Region. The architecture’s scalability and resilience handled this growth with no modification to architectural components. In 12 months of production within only a single AWS Region, around 200 million lambda invocations successfully automated job submissions, with less than 250 errors (0.000125% error rate). These Lambda functions were triggered by over 20 million SNS messages, without incidence of error.

Celeste manages bursty workflows that are characterized by sudden spikes in computational demand, ensuring rapid processing of large genomic datasets. Additionally, cloud resources address the need for specialized compute hardware (FPGA or GPU), which is often a prerequisite for the intensive data analysis involved in NGS. Lastly, when preparing for large scale projects, it is often impossible to precisely predict the required storage and compute resources, especially as project complexity causes timelines to change. Because of this, cloud environments, which enable a very wide range of workloads are appealing solutions. Celeste provides storage flexibility, a vital feature given the massive amounts of data generated. This adaptability allows for scalable storage solutions that can expand or contract in response to fluctuating data volumes, thereby optimizing costs and resource utilization. Together, these features make Celeste a robust tool for overcoming the complexities inherent in NGS workflows. We have open-sourced this tool so that other groups can benefit from our experience.

The initial pipeline was implemented using DRAGEN, which offers a number of advantages. We made the decision to prioritize the detection of low allele fraction variants because these variants may be important clinically and their prevalence in disease may be important subjects for research in the future.

The learning curve for cloud technologies is steep, due to the breadth of knowledge required. Cloud providers offer a wide array of rapidly evolving services, each with its own features and use cases. Security and cost must be constantly monitored, along with following best practices that are scalable, resilient, and efficient. Because of this, cloud teams must be exceptionally diverse in terms of their skillset. Addressing these challenges often requires a mix of formal education, self-study, hands-on practice, and staying actively engaged with the cloud computing community to remain current with trends and best practices.

Cloud architectures offer a large number of advantages for NGS pipelines. We hope the open-sourced Celeste Cloud Formation templates can serve as a model to other groups seeking to take advantage of cloud resources.

## Conclusions

Celeste offers a flexible architecture for implementing analysis pipelines for genomic medicine. It has been tested at a very large scale and is capable of handling a wide range of workflows, including alignment and variant calling with DRAGEN, QC metric generation, variant liftover, concordance checking and variant annotation, in support of projects like *All of Us*^*7*^, Neurocare^30^ and Epilepsy^31^.

## Methods

In a method similar to many AWS QuickStart repositories, we take advantage of tools such as GitHub, CodePipeline, CodeBuild, and CloudFormation to build, version, and release our cloud infrastructure along with all software components. We utilize TaskCat and CI/CD pipelines within the AWS Developer Tools to build an ephemeral mock-up of our cloud infrastructure in a test VPC, before submitting a status report and merging those changes to our development environment. Releases to our staging and production environments happen in much the same way, with the addition of deployer control and smoke test job submissions to validate the results of the variant-calling pipeline.

The pipeline was benchmarked on control samples described previously^6^. A specialized sample (Tru-Q 1 (5% Tier) Reference Standard) from Horizon Discovery (Cambridge, UK)^32^ was used to assess the limit of detection.

### Components of the Initial pipeline

The initial pipeline infrastructure utilizes CloudFormation templates to provision necessary components like SNS topics, Lambda functions, Step Functions, and AWS Batch resources. Event-driven architecture elements consist of new object notifications (PUT, POST, COPY, or multipart uploads) and specialized Lambda functions (Scandium, VerifyBamID, Alignstats^33^, S3Tag, Liftover Intersect, Liftover Preprocessing, Intervar^34^, Stargazer^35^. Batch architecture elements include job definitions, compute environments, and job queues for different DRAGEN versions, reports jobs, high-memory jobs and Intervar. DRAGEN software is distributed by Illumina in an AMI, and we have additionally created an AMI for QC jobs that follow the primary and secondary analysis. Containers for further analyses (cassandra, Mpileup, Intervar, alignstats0.9, liftover, optitype, scandium1.4.2, stargazer1.0.9, verifybamid1.1.3) are managed using Docker and stored in ECR.

### Merge workflows

Typical NGS workflows employ multiple sequencing events for a single sample, often containing a calibration run, followed by additional sequencing events to reach full coverage. Celeste is designed to handle these merge workflows, processing both single and multiple sequencing events.

### DRAGEN versions

Multiple DRAGEN versions were used during benchmarking, most importantly v3.3.7 for evaluating low allele fraction parameter sets and v3.4.12 for generating the IDE validation data. *All of Us* initially began production using DRAGEN 3.4.12 and later moved to 3.7.8. All samples were reprocessed to match the 3.7.8 version.

### DRAGEN parameter tuning

The DRAGEN team performed a grid-search over the parameter space in order to find parameters that allowed the low allele fraction variants to be called in the control sample, while minimizing the overall change in the f-measure.

## Data Availability

The public release of the Celeste infrastructure is available here: https://github.com/NooraSiddiqui/Celeste. Data produced by the All of Us Research Program can be accessed by registered researchers in the All of Us research workbench here: https://workbench.researchallofus.org/login.

https://github.com/NooraSiddiqui/Celeste

## Data and Code availability

## Declarations

### Ethics approval and consent to participate

Not applicable

### Consent for publication

Not applicable

### Availability of data and materials

The public release of the Celeste infrastructure is available here: https://github.com/NooraSiddiqui/Celeste. Data produced by the *All of Us* Research Program can be accessed by registered researchers in the *All of Us* research workbench here: https://workbench.researchallofus.org/login.

### Authors’ contributions

NS and EV designed the work. EV, RAG, NJL and RM conceived of the work. NS, BL, VY, SM and SC created new software used in the work. NS, JF, ZK, SEK, QW, KW, JM, CK, and MG analyzed and interpreted the data. NJL, RAG and EV aquired the data. NS and EV wrote the manuscript. All authors reviewed the manuscript.

## Availability and Requirements

Project name: Celeste

Project home page: https://github.com/NooraSiddiqui/Celeste

Operating system(s): e.g. Platform independent

Programming language: Python, CloudFormation

License: https://opensource.org/license/mit

Any restrictions to use by non-academics: None

## Acknowledgements

The All of Us Research Program would not be possible without the partnership of its participants. To learn more about the All of Us Research Program’s research data repository, please visit www.researchallofus.org/.

